# Identifying the neurodevelopmental and psychiatric signatures of genomic disorders associated with intellectual disability

**DOI:** 10.1101/2022.12.16.22283581

**Authors:** Nicholas A Donnelly, Adam C Cunningham, Matthew Bracher-Smith, Samuel Chawner, Jan Stochl, Tamsin Ford, F Lucy Raymond, Valentina Escott-Price, Marianne BM van den Bree

## Abstract

**Introduction:** Genomic conditions can be associated with developmental delay, intellectual disability and physical and mental health symptoms, but are individually rare and variable, which limits the use of standard clinical guidelines. A simple screening tool to identify young people with genetic conditions associated with neurodevelopmental disorders (ND-GC) who could benefit from further support would be of considerable value. We used machine learn approaches to address this question.

**Methods:** A total of 489 individuals were included: 376 with a ND-GC, mean age=9.33, 63% male) and 113 unaffected siblings; mean age=10.35, 50% male). Primary carers completed detailed assessments of behavioural, neurodevelopmental and psychiatric symptoms and physical health conditions. Machine learning techniques (elastic net regression, random forests, support vector machines and artificial neural networks) were used to develop classifiers of ND-GC status using a limited set of variables. Exploratory Graph Analysis was used to understand associations within the final variable set.

**Results:** We identified a set of 30 variables best discriminating between ND-GC carriers and control individuals, which formed 4 dimensions: Anxiety, Motor Development, Insomnia and Depression. All methods showed high discrimination accuracy with Linear Support Vector machines outperforming other methods (AUROC between 0.959 and 0.971).

**Conclusions:** In this study we developed models that identified a compact set of psychiatric and physical health measures that differentiate individuals with a ND-GC from controls and highlight the structure within these measures. This work is a step toward developing of a screening instrument to select young people with ND-GCs who might benefit from further specialist assessment.

## Introduction

Up to 20% of patients with a neurodevelopmental disorder have an identifiable genomic condition (1–4). Such conditions include copy number variants, single nucleotide variants and aneuploidies, which we collectively call neurodevelopmental genomic conditions (ND- GC). ND-GCs have been associated with schizophrenia (5), attention deficit hyperactivity disorder (ADHD), autism spectrum disorder (ASD) (6), and intellectual disability (ID) (7).

The clinical presentation of ND-GCs is variable and complex. For example, children with 22q11.2 deletion syndrome, a disorder caused by a deletion in the q11 region of chromosome 22, have a high risk of developmental delay and intellectual disability (8), seizures (57%) (9), motor coordination problems (81%) (10), sleep disturbances (60%) (11) and psychiatric disorders (12). Such complex presentation is not unique to 22q11.2 deletion but is typical for many ND-GCs (13), as is incomplete and variable penetrance (14,15).

It is therefore extremely important for families of a child with an ND-GC to be informed about the impact that the variant may have on their child’s development, so that they can obtain the best possible support. Additionally, clinicians, such as psychiatrists in CAMHS and community learning disability services, who care for affected children after diagnosis are challenged by complex presentations where symptoms which may require input from multiple clinical specialities are present.

This problem can be exacerbated by variability in the conditions that present in children with a ND-GCs, which may not follow the expected symptom patterns based on research from non-genotyped populations. For example, we have observed that children with 22q11.2 deletion and ADHD are much more likely to be affected with an inattentive subtype than the children with idiopathic ADHD (16). A clinician who is unaware of this may be less likely to diagnose ADHD, meaning that the child misses beneficial treatment. Diagnostic overshadowing may also take place, a well-recognised phenomenon where difficulties that are experienced by a child with a genomic disorder are interpreted as wholly due to ID (17–19). This can reduce the chance for referral to appropriate services and access to appropriate treatment (20,21).

One solution to these problems would be to identify patterns of neurodevelopmental and physical health symptoms that are most associated with carrying a ND-GCs, to stratify affected patients for graded approaches to investigation and treatment. In the present study, we identify those symptoms that most robustly differentiate between children with ND-GCs and typically developing control siblings, and analyse whether these symptoms form broader symptom domains, using a large sample of children with a wide range of ND-GCs and deep physical and mental health phenotyping.

## Methods and Materials

### Participants

We defined ND-GCs as conditions associated with increased risk of neurodevelopmental symptoms (22) and caused by a genetic variant which was either pathogenic or likely pathogenic, according to American College of Medical Genetics and Genomics guidance (23). We aimed to recruit a population of participants with a range of ND-GCs that represented a “snapshot” of presentations to UK CAMHS, intellectual disability, clinical genetics or community paediatrics clinics.

Families of children with ND-GCs were recruited through UK Medical Genetics clinics, word of mouth and the charities UNIQUE (https://rarechromo.org) and MaxAppeal! (https://www.maxappeal.org.uk), as part of ongoing cohort studies at Cardiff University including the ECHO study (https://www.cardiff.ac.uk/cy/centre-neuropsychiatric-genetics-genomics/research/themes/developmental-psychiatry/copy-number-variant-research-group) and the IMAGINE study (https://imagine-id.org) (13,22).

In total 589 individuals (441 individuals with a ND-GC and 148 unaffected control siblings) were included in the study, from whom data from 489 individuals was included in our machine learning analysis after initial data preparation (**Supplementary Methods**). Our sample size was the maximum number of participants in our dataset who had all the required variables.

Informed, written consent was obtained prior to recruitment from the carers of participants and recruitment was carried out in agreement with protocols approved by relevant NHS and university research ethics committees. Individual ND-GC genotypes were established from medical records and in-house genotyping at the Cardiff University MRC Centre for Neuropsychiatric Genetics and Genomics using microarray analysis. Participant genotypes are shown in Supplementary Table 1.

### Assessments

Primary carers of participants completed a battery of assessments to collect comprehensive information on physical and mental health problems through semi-structured interview with trained research staff and questionnaires. Assessments were carried out between January 2011 and December 2019. Our goal was to generate a set of items that could be easily and conveniently completed by a carer or community clinician either on paper or online.

Therefore, measures which involved complex or invasive testing, such as cognition or blood tests, were not included in our analysis.

Psychiatric symptoms were measured using the Child and Adolescent Psychiatric Assessment (CAPA, (24)), Strengths and Difficulties Questionnaire (SDQ, (25)) and the Social Communication Questionnaire (SCQ, (26)). The CAPA assesses domains including ADHD, anxiety disorders, oppositional defiant disorder, obsessive compulsive disorder, psychosis and psychotic experiences, tic disorders, mood disorders, and substance abuse. The SDQ is a dimensional measure of psychopathology that includes measures of hyperactivity, emotional problems, peer problems, and prosocial behaviour. The SCQ measures ASD-associated symptoms and was used as the CAPA and SDQ lack of coverage of ASD symptoms.

Additionally, as mounting evidence indicates difficulties with coordinated movement are an important symptom in individuals with ND-GCs (10,13,27,28), we assessed coordination using the developmental coordination questionnaire (DCDQ, (29)).

Information about physical health problems and development was collected through a questionnaire including questions asking about presence or absence of heart problems, seizures, musculoskeletal problems, and respiratory problems.

### Statistical Analysis and Data Availability

All statistical analysis was carried out in R version 4.2.1 (30). An overview of the analysis workflow is presented in Figure 1. Code used in the project is provided in a GitHub repository https://github.com/NADonnelly/nd_cnv_ml and fitted models are presented as an interactive Shiny app: https://nadonnelly.shinyapps.io/cnv_ml_app/. Data from the IMAGINE study is available via the IMAGINE ID study website: https://imagine-id.org/healthcare-professionals/datasharing/. Analysis is reported in line with the TRIPOD guidelines, Supplementary Table 2 (31). An early version of this manuscript was deposited as a preprint:.

**Figure 1.**
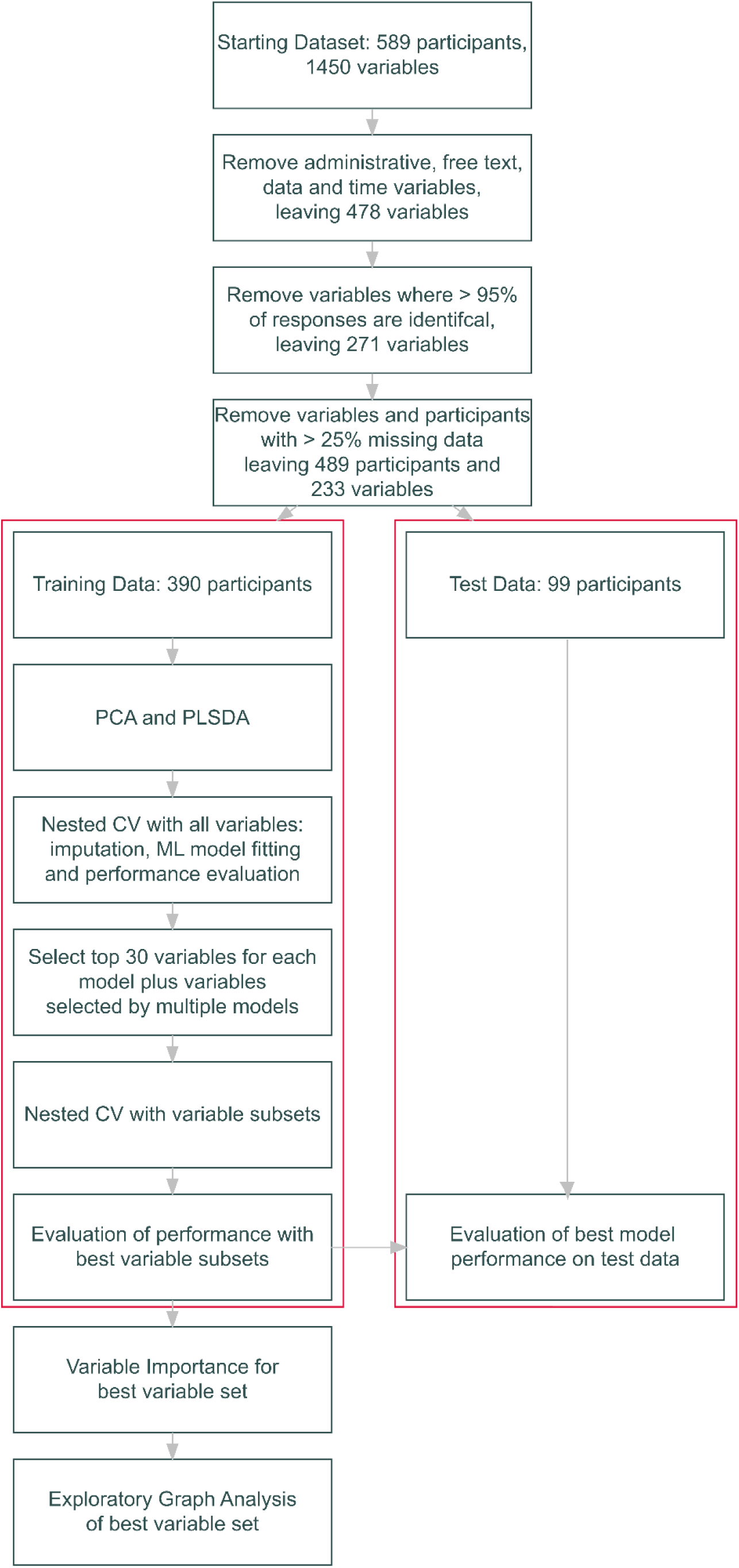
Flowchart of analysis workflow including variable and participant selection and machine learning model fitting. CV: Cross-validation; ML: Machine Learning; PCA: Principal Components Analysis; PLSDA: Partial Least Squares Discriminant Analysis

### Dimensional Structure Assessment

We applied principal components analysis (PCA) followed by partial least squares discriminant analysis (PLSDA, where the outcome was ND-GC status) to explore the dimensional structure of our dataset, using the *mixOmics* package (32). A cross-validation process was used find the optimal number of components and variables for the PLSDA (**Supplementary Methods**).

### Machine Learning (ML) Model Fitting

We prepared our data for ML model fitting by splitting participants into a training dataset of 390 (80% of the dataset) and a test set of 99 (20% of the dataset), stratifying by ND-GC status.

Our outcome was binary classification of ND-GC status (carrier vs control), and we evaluated model performance using the area under the receiver operator characteristic curve (AUROC). We used elastic net regression (using the *glmnet* package (33)), random forests (using the *Ranger* package (34)), linear support vector machines (SVMs, using the *kernlab* package (35)) and single layer artificial neural networks (using the *nnet* package (36)) to create models capturing linear and non-linear relationships.

Models were fit using nested cross-validation (CV), with 20 outer folds and 20 inner folds. Outer folds were generated by splitting the data into 5 folds, repeated 4 times. Inner folds were generated from the outer fold analysis set using bootstrapping with replacement. Within each outer fold missing data was imputed using bagged tree models (37) and the same model was used to impute missing data in the analysis set.

Grid search (30 elements) was used to optimise hyperparameters for ML models across inner folds. Model performance was evaluated by fitting the model with the best performing set of hyperparameters in the inner fold data to the (previously unseen) outer fold assessment dataset. This process was then repeated for all outer folds.

Following nested CV, we selected models with the highest AUROC, and evaluated the importance of all included variables for model prediction using permutation testing (38). We selected the top 30 variables for all ML models and generated two further variable sets: all variables which were included in the top 30 most important for more than one ML model, and those variables included in the top 30 for at least 3 models, to give a total of 6 sets of variables.

We extracted 30 variables for each model because we wanted to achieve a balance between accurate prediction, including a wide set of variables for exploration of dimensional structure and limiting the number of items to that which could be realistically completed by young people’s carers and/ or clinicians.

We repeated our nested CV process, using the same ML models using the 6 sets of most-predictive variables, giving a total of 24 combinations of models and predictor variables, selecting the best performing combinations of variables and ML model, based on AUROC.

We evaluated the performance of the final models using the held-out training data. Missing data in the test dataset was imputed using a model fit to the full training dataset, and the ND- GC status of each participant in the test dataset was predicted using the best ML models.

Model performance was evaluated by drawing 2000 bootstrap samples from the test dataset and estimating performance (AUROC and mean log loss) for the bootstrap sample. This produced a distribution of values from which a median value and a 95% confidence interval were calculated.

Model calibration i.e., the relationship between true and model-predicted probability of ND- GC status, was estimated by binning model predictions by predicted probability of ND-GC status and plotting this against true ND-GC status. Model performance was also estimated for male and female participants separately, and after binning participants by age quintile.

The importance of each variable in the best fitting model was evaluated using a permutation-based approach, as above.

The optimal threshold for converting model predicted probability of ND-GC status into a binary classification was estimated by finding the threshold which maximised the j-index (sensitivity + specificity – 1, (39)).

### Exploratory Graph Analysis

Bootstrap Exploratory Graph Analysis (EGA) was used to investigate the dimensional structure of the best performing variable set. EGA has been shown to be as accurate or more accurate than traditional factor analytic methods such as parallel analysis (40,41). Bootstrap EGA estimates and evaluates dimensional structure in a set of variables by first applying a network estimation method (*EBICglasso* as applied using the *qgraph* package (42)), followed by a community detection algorithm for weighted networks (Walktrap community detection algorithm (43)). Non-parametric bootstrapping is then used to generate bootstrap samples (n = 10,000) from the input dataset, and EGA was applied to each replicate sample to form a sampling distribution from which the median value of each edge across the replicate networks, resulting in a single network. The stability of the network can be assessed by measuring the proportion of bootstrapped networks where a given variable is included in each putative dimension (41), and the number of variables included can be adjusted to improve the stability of dimension representations. We therefore fit an EGA model to a full set of variables, then repeated the analysis with the variables with the most consistent relationship to our dimensions (item stability > 0.75; this left 20 variables), generating a stable and consistent EGA model.

To provide an additional assessment of the fit of the proposed dimensional structure to the data, confirmatory factor analysis was carried out on the typical dimension structure identified by bootstrap EGA, with fit assessed using CFI and RMSEA.

Finally, we repeated the above model fitting processing using the most important variables in each of the identified four dimensions identified by EGA.

## Results

### Study Participant Characteristics

Characteristics of study participants are given in Supplementary Table 3 and genotypes in Supplementary Table 1. Individuals with an ND-GC were approximately a year younger than controls and there was a higher proportion of males in the ND-GC group. Compared to families where both a control and a ND-GC carrier took park, families where just a ND-GC carrier took part had lower parental educational level and income, and there were fewer participants of European ancestry; the discrepancy between ND-GC carriers and control individuals was due to most ND-GC carriers not having a sibling included in the study (59%).

### Partial Least Squares Analysis

We applied principal components analysis (PCA) and partial least squares discriminant analysis (PLSDA) to our full set of variables 233 for the 390 participants in our training dataset to describe the dimensional structure of our variables. This analysis indicated that one component explained a particularly large proportion of the variance (16.6%, Supplementary Figure 1), with the second and third components (5.7 and 3.9% respectively) also providing useful explanation of variation. We applied PLSDA to our dataset, a supervised dimension reduction method which focusses on discrimination between groups. We found that 3 components provided optimal discrimination between groups, with 140, 220 and 230 variables selected for each of the three components, respectively (Supplementary Figure 1). This analysis indicated that it was possible to identify ND-GC carriers from controls using our dataset, with ND-GC carriers having higher scores on component 1. Some individuals with a ND-GC showed similar profiles to controls and likely represent participants with a ND-GC that are relatively mildly affected; some controls showed profiles more like those with ND-GCs, reflecting individuals in the control sample with elevated difficulties across the measured domains.

**Supplementary Figure 1.**
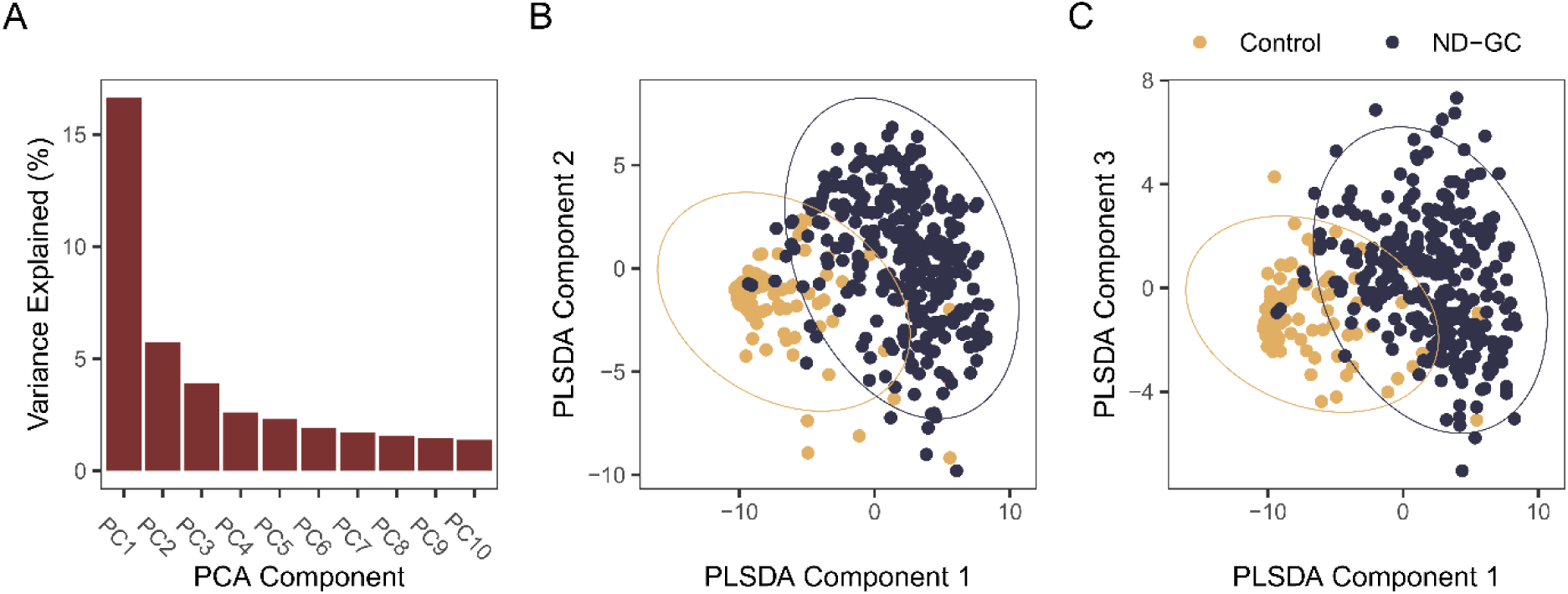
PCA and PLSDA. A: Variance explained by the first 10 principal components of all 233 variables in 390 participants in the training dataset. One component explains a particularly large proportion of variance (16.6%). B: scatter plot of all participants by the first two PLSA components, with 95% confidence ellipse for each class. C: as B, for PLS components 1 and 3.

However, this analysis still selected large numbers of variables. We applied machine learning approaches to develop classification models with an optimally predictive subset of variables.

### Developing machine learning models

We developed machine learning models to classify individuals by ND-GC status, including artificial neural networks (ANN), linear support vector machines (SVM), penalised logistic regression (LR) and random forest classifiers, using our full training set of 233 variables and 390 participants, with nested cross validation (CV). After nested CV, all models performed well at distinguishing between individuals in the training data set with a ND-GC and controls, with median AUROCs ≥ 0.9 in all cases (Supplementary Table 4). The SVM performed best, with an overall median AUROC of 0.936. The random forest and penalised logistic regression models did not perform significantly worse than the SVM, but the performance of the ANN was significantly poorer (AUROC difference = -0.036, 95% credible interval of difference [-0.047, -0.025]).

### Predictive performance with optimised variable sets

We repeated model fitting using nested cross validation using the sets of variables selected as being most important to the models fit to the full set of variables (determined using permutation testing). Results were similar across multiple models and variable sets (Figure 2A, Supplementary Table 5). We selected the “SVM” variable set for further analysis as this set appeared to produce both the single best classification performance (the combination of the linear SVM model and SVM variables) and the best performance across multiple model types.

**Figure 2.**
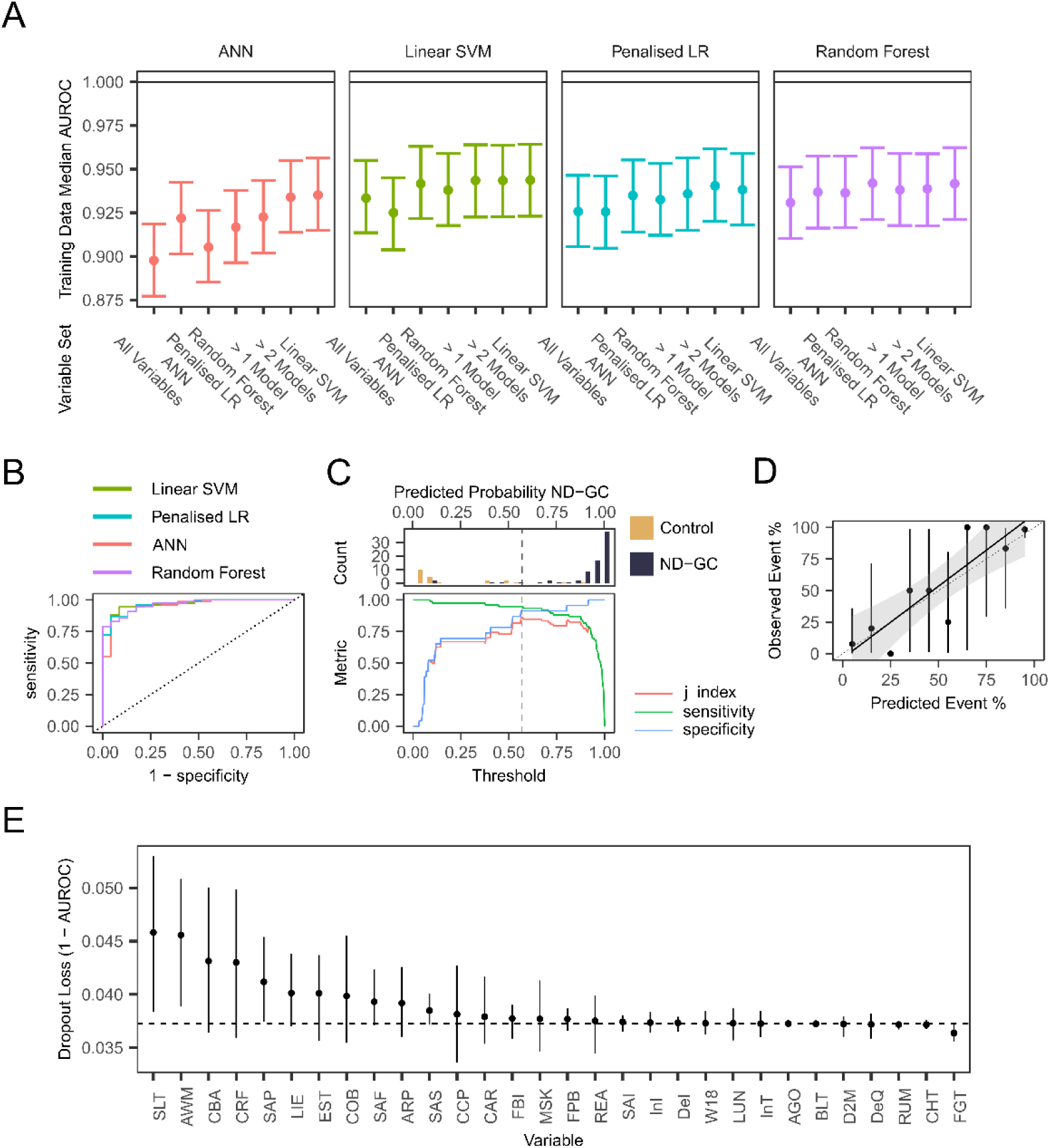
Performance of final models on test data. A: Plot of performance (AUROC) of four machine learning models (ANN = Artificial Neural Network, Penalised LR = Penalised Logistic Regression, Linear SVM = Linear Support Vector Machine fit to 7 difference variable sets (All Variables = All 233 variables; ANN = the top 30 most important variables identified by an ANN fit to all variables; Penalized LR = the top 30 most important variables identified by a penalized logistic regression fit to all variables; Random Forest = the top 30 most important variables identified by a random forest model fit to all variables; > 1 Model = variables identified as being in the top 30 most important variables by more than one ML model; > 2 Models = variables identified as being in the top 30 most important variables by more than two ML models; Linear SVM = the top 30 most important variables identified by a linear SVM fit to all variables. Points show the median posterior AUROC, error bars show the 95% credible interval of the AUROC. B: Receiver-operator characteristic curves for the 4 machine learning models, using the 30 variables from the linear SVM dataset). C: Top – histogram of predicted probability of ND- GC status in the 99 participants in our testing dataset using the best performing Linear SVM model; Bottom – plots of sensitivity, specificity of model classification performance at different thresholds for categorising a predicted probability into control or ND-GC. Optimal performance, as indexed by the j-index (sensitivity + specificity – 1) occurred at a threshold of 0.57. D: Calibration plot for the best performing linear SVM. The sensitivity plot compares the predicted and true probability of participants being ND-GC carriers across 10 bins of predicted probability. Points are performance in each decile, vertical lines show 95% confidence intervals, thick diagonal linear shows a linear model fit to the data, with the shade area showing the 95% confidence interval of the linear model. A perfectly performing model would following the diagonal dashed line. E: Variable importance for the best fitting model. Mean dropout loss is the mean change in model AUROC after a given variable is permuted (repeated 500 times). Horizontal line indicates (1 – AUROC) of the full model; therefore, variables with mean values above this line have a negative impact on model fit when permuted. Variable definitions: see table 6 for full definitions; SLT: Ever had speech therapy; AWM: Invented words, odd indirect, metaphorical ways; CBA: catches a small ball thrown from 6-8ft; CRF: runs as fast and easily as other children; SAP: Physical symptoms on separation intensity; LIE: Lying; EST: educationally statemented; COB: can organise her body to do a planned motor activity; SAF: Separation Anxiety if not co-sleeping with a carer; ARP: say the same thing over and over; SAS: Avoidance of sleeping away from family; CCP: cuts pictures and shapes accurately; CAR: heart problems; FBI: Fear of blood/injection; MSK: skeletal or muscular problems; FPB: Fear of activities in public avoidance; REA behind in reading; SAI: Separation worries/anxiety; InI: Initial insomnia intensity; DeI: Episode of depressed mood intensity; W18: walking by 18 months; LUN: other problems with airways/lungs; AGO: Agoraphobia; InT: Insomnia intensity; BLT: Often blurts out answers to questions; D2M: Period of 2 continuous months without depressed mood in last year; DeQ: Distinct quality of depressed mood; RUM: Rumination; CHT: Cheating; FGT: Forgetful in daily activities

We then fit the best performing models to our held-out test set of data from 99 participants. Classification performance with this test dataset was (Figure 2B, Table 1). The best performing model was an SVM, achieving an AUROC of 0.971 (95% CI [0.942, 0.997]) with a mean log loss of 0.197 (95% CI [0.110, 0.286]). This model correctly classified 72/76 ND-GC carriers (94.75%) and 18/23 controls (78.3%). Performance of other models was not significantly poorer than the SVM. The optimal probability for classifying a participant as a ND-GC carrier, the point at which the j-index is maximised, was 0.57 (Figure 2C).

**Table 1 Caption:**
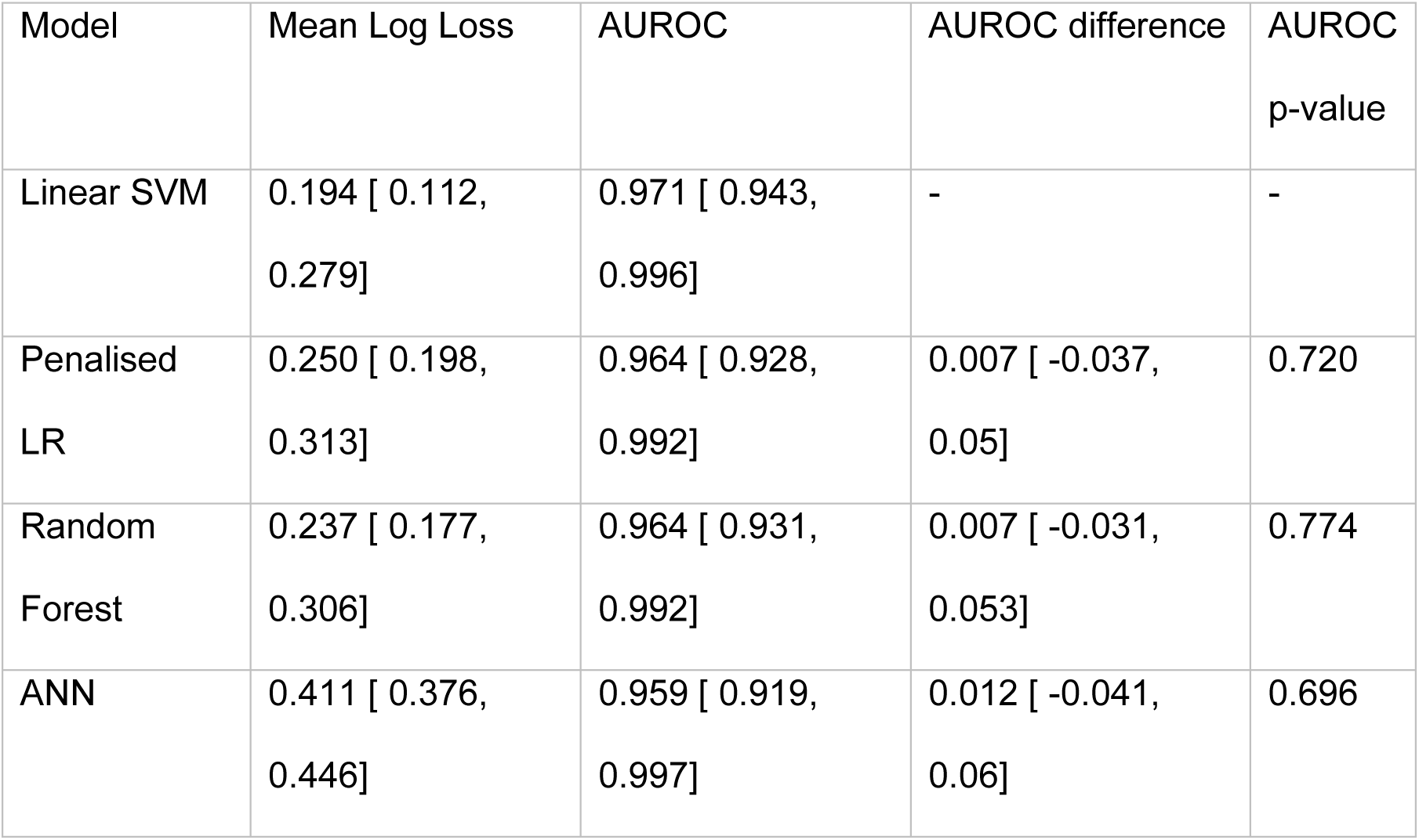
Final model performance on held-out test dataset. Values shown are bootstrapped performance and the 95% confidence interval of the measure (Mean Log Loss and AUROC), and difference in AUROC between the linear SVM and the other models, with its 95% confidence interval, and the p-value of the AUROC difference

| Model | Mean Log Loss | AUROC | AUROC difference | AUROC<br>p-value |
| --- | --- | --- | --- | --- |
| Linear SVM | 0.194 [ 0.112,<br>0.279] | 0.971 [ 0.943,<br>0.996] | - | - |
| Penalised<br>LR | 0.250 [ 0.198,<br>0.313] | 0.964 [ 0.928,<br>0.992] | 0.007 [ -0.037,<br>0.05] | 0.720 |
| Random<br>Forest | 0.237 [ 0.177,<br>0.306] | 0.964 [ 0.931,<br>0.992] | 0.007 [ -0.031,<br>0.053] | 0.774 |
| ANN | 0.411 [ 0.376,<br>0.446] | 0.959 [ 0.919,<br>0.997] | 0.012 [ -0.041,<br>0.06] | 0.696 |

We investigated whether classification performance varied over participant age or between genders. Performance appeared to be marginally higher in male than female participants, but the difference was small, and there did not appear to be consistent differences in performance across participant ages, although our sample was mostly of younger participants (Supplementary Table 6).

Analysis of model calibration demonstrated some miss-calibration between predicted and actual probabilities, with the model having some tendency to given higher-than-optimal predicted probabilities of ND-GC status at higher predicted probabilities (Figure 2D).

We investigated variable importance in our best performing model (Figure 2E). This demonstrated that a subset of variables appeared to have a particularly large importance to the model. We next investigated whether there was a dimensional structure within our variable set that could be used to understand the predictors of ND-GC status.

### Underlying dimensional structure of selected variables

We next investigated an underlying structure of the variables included using an exploratory graph analysis (EGA). The 30 variables used were the optimised variable set of the best performing SVM model, determined using permutation testing. These variables included items from the Developmental Coordination Disorder Questionnaire, Social Communication Questionnaire, Child and Adolescent Psychiatric Assessment and the Health and Development Questionnaire.

EGA fit to the most stable set of variables (20 variables were included in the final EGA model) revealed that the variables formed a structure consisting of 4 dimensions: 1: Anxiety (predominately separation anxiety and agoraphobia/fear of public places); 2: Developmental Milestones and Motor Co-ordination; 3: Insomnia and 4: Depression (Figure 3, Supplementary Table 7).

**Figure 3.**
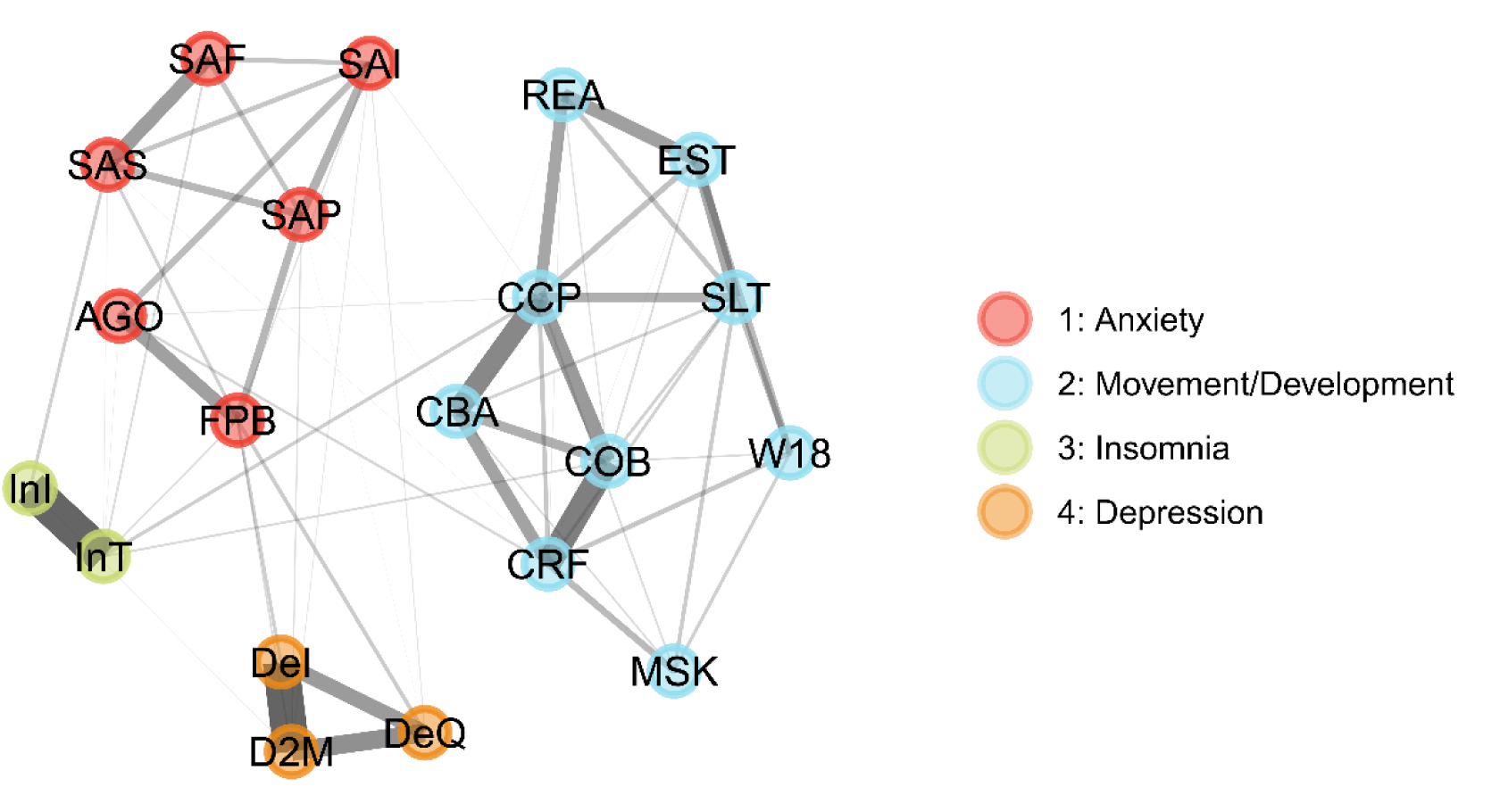
Exploratory Graph Analysis. The graph shows correlations between variables (notes) as lines, where line thickness represents correlation strength. Nodes are coloured by the putative dimensions they are assigned to by the Bootstrapped EGA algorithm. Variable Definitions: See Table 6 for full variable definitions; AGO: Agoraphobia; CBA: catches a small ball thrown from 6-8ft; CCP: cuts pictures and shapes accurately; COB: can organise her body to do a planned motor activity; CRF: runs as fast and easily as other children; D2M: Period of 2 continuous months without depressed mood in last year; DeI: Episode of depressed mood intensity; DeQ: Distinct quality of depressed mood intensity; EST: educationally statemented; FPB: Fear of activities in public avoidance; InI: Initial insomnia intensity; InT: Insomnia intensity; MSK: skeletal or muscular problems; REA: Is your child behind in reading; SAF: Separation Anxiety; SAI: Separation worries/anxiety intensity; SAP: Physical symptoms of separation intensity; SAS: Avoidance of sleeping away from family intensity; SLT: Has your child had speech therapy; W18: Did your child walk by 18 months

Confirmatory factor analysis based on this four-dimension structure demonstrated that the 4- factor structure fit with RMSEA of 0.052 and CFI of 0.934, indicating a reasonable fit to the data.

Finally, we investigated if the variable domains identified through EGA could be used to develop a further reduced set of variables for use in a ML model; although a 30-item scale could be realistically used in a clinical setting, a much shorter screener could be useful in busy clinical environments. We therefore selected the variable in each dimension with the highest variable importance from our 30 item SVM ML model and fit a linear SVM model to our training data, using these 4 variables. A linear SVM fit to 4 variables (DeI [depression intensity], SAP [physical symptoms of separation anxiety], InI [initial insomnia], SLT [history of speech and language therapy]) had an AUROC = 0.955 [0.914, 0.993] and mean log loss = 0.253 [0.203, 0.308], with 70/76 participants with an ND-GC being correctly classified (92.1%), and 22/23 control participants classified correctly (95.7%). This performance was lower than the full 30 variable model, but still indicative of high absolute classification performance.

## Discussion

### Main findings

In this study we demonstrate the potential of using machine learning to identify key variables where individuals with genetic conditions associated with intellectual disability and neurodevelopmental disorders differ from unaffected control individuals, based on a limited set of psychiatric, behavioural and physical health related variables, in the absence of biochemical, genetic, IQ or neurocognitive data. Using an SVM classifier, we were able to classify individuals with an ND-GC with excellent performance, achieving an AUROC of 0.971. We identified 4 dimensions in our variable set that appeared to be most relevant to identifying individuals with an ND-GC, namely, development/health, anxiety, insomnia and depression.

### Relationship to previous studies

Previous studies have described the high rates, and complex presentations, of psychiatric and neurodevelopmental difficulties in children with ND-GCs (8,12,13,22,44). ND-GCs are associated with a wide range of health outcomes (15), along with multimorbidity later in life (45), and are highly enriched in the population with developmental delay/intellectual disability (1,3,4,46). However, not all individuals with a ND-GC will meet diagnostic criteria for specific psychiatric disorders (47). We attempted to address this by not including diagnostic status in our classification models, only symptom scores; the highly accurate classification we were able to achieve supports the idea that profiles of symptoms are most informative when identifying areas of relative difficulty or strength in individuals with ND-GCs.

We identified 4 underlying dimensions in our final set of 30 variables. These dimensions identify potential key phenotypic areas where individuals with ND-GCs differ from controls: anxiety (particularly separation anxiety), motor skills and development, insomnia, and depression, as well as suggesting that other domains, such as difficulties with conduct or hyperactivity, may be less discriminating. The identified dimensions map onto areas of difficulty elucidated in previous studies (11,27,47–50), and highlight that specific symptoms may be particularly informative about ND-GC status, including initial insomnia, intensity of depressive symptoms, physical symptoms of separation anxiety, and having a history of speech and language therapy.

Clinical care pathways may be enhanced by focusing more on the areas identified as key dimensions by our analysis if further research demonstrates that they are areas that predict longer term difficulties for children with ND-GCs. It will also be important to take the items identified and work with parents and clinicians to optimise the wording and content of any items that could be used in a screening test derived from our analysis. For example, one highly predictive item refers to a history of speech and language therapy. As ND-GC carriers can struggle to access therapies in a timely fashion, this item might miss individuals who might have needed speech and language therapy, but not been able to access it; therefore, asking about relative difficulties with speech and language may be more informative.

### Strength and limitations

This is the largest study of its kind to investigate the possibility of differentiation between individuals with a broad range of ND-GCs and controls based solely on psychiatric and health phenotypes using machine learning models. We were able to produce a model with very high AUROC, which appeared to perform well across a range of relevant ages, and in both males and females.

However, while including a very broad range of genomic disorders provided a more representative sample of those variants seen by clinical services, it may have increased the noise and variability in symptoms. Our sample was also unbalanced, in that there were a larger number of individuals with a ND-GC than controls, because not all families with a child with an ND-GC had an unaffected sibling of a similar age. This can affect model performance, as most techniques work best in balanced samples.

Our initial partial least squares discriminant analysis on our full dataset of phenotypic and psychiatric information indicated that young people with an ND-GC and control individuals lie on a spectrum of symptoms, and that while it is possible to distinguish between the two groups based on psychiatric, behavioural and health information, there remain some individuals with a ND-GC who have profiles that are very similar to unaffected individuals.

This highlights the wide variety of phenotypic expression that is seen within individuals with ND-GCs, which will impose limits on the performance of any classification algorithm.

Additionally, ascertainment bias may affect our results. Developmental delay is a major reason for referral for genetic testing in the UK, and it is likely that our sample has a preponderance to include those individuals with ND-GCs who are on the more severe end of the phenotypic presentation, and as such it may be the case that the common dimensional structure we identify as being associated with ND-GC carriage may be applicable only to relatively more severe difficulties, rather than the phenotype of the entire population of ND- GC carriers.

Our machine learning models and EGA would be strengthened by measuring performance and performing confirmatory factor analysis using an independent sample. Future studies which combine measurement of most differentiating variables and longer-term follow-up of psychiatric and health outcomes would allow the predictive accuracy of our model to be evaluated.

We considered the role of decision curve analysis in our study, as this approach has been recommended in studies of prediction models (51). However, such calculations rely on samples being drawn from a population comparable to the clinical population. Our study sample was drawn from a cohort explicitly required to be ND-GC carriers (or sibling controls). Therefore, such an analysis is not applicable to our study. However, it should be performed in a future study validating our model in a broader population.

Despite these limitations, it is important to better understand the difficulties faced by this group of patients as they make up a significant proportion of those presenting to intellectual disability services and clinicians often lack complete information on prognosis for patients with ND-GCs. This study highlights areas of difficulties for those children who may most need further support, which may warrant further research and may be targets for individualised interventions.

## Conclusions

We demonstrate that it is possible to accurately detect individuals with ND-GCs associated with neurodevelopmental disorders and intellectual disability based on a limited set of psychiatric and health variables which could form the basis for clinical screening instruments. We highlight that separation anxiety, development of motor skills and speech, insomnia and depression are important areas where children with ND-GCs differ from control individuals. Future research should investigate these areas in more detail so that targeted interventions can be developed.

## Data Availability

Code used in the project is provided in a GitHub repository https://github.com/NADonnelly/nd_cnv_ml and fitted models are presented as an interactive Shiny app: https://nadonnelly.shinyapps.io/cnv_ml_app/. Data from the IMAGINE study is available via the IMAGINE ID study website: https://imagine-id.org/healthcare-professionals/datasharing/.

https://nadonnelly.shinyapps.io/cnv_ml_app/

https://imagine-id.org/healthcare-professionals/datasharing/

## Author Contributions

Nicholas Donnelly: Methodology, Software, Formal analysis, Data Curation, Visualisation, Writing – Original Draft, Writing – Review & Editing

Adam Cunningham: Methodology, Software, Formal analysis, Data Curation Matthew Bracher-Smith: Methodology

Samuel Chawner: Conceptualisation, Investigation, Writing – Review & Editing, Funding acquisition

Jan Stochl: Methodology, Writing - Review & Editing

Tamsin Ford: Writing – Review & Editing, Funding acquisition

F Lucy Raymond: Conceptualisation, Writing – Review & Editing, Funding acquisition

Valentina Escott-Price: Conceptualisation, Methodology, Writing – Review & Editing, Funding acquisition

Marianne BM van den Bree: Conceptualisation, Writing – Review & Editing, Funding acquisition, Project administration, Supervision

## Ethical standards

The authors assert that all procedures contributing to this work comply with the ethical standards of the relevant national and institutional committees on human experimentation and with the Helsinki Declaration of 1975, as revised in 2008.

## Acknowledgements

We are extremely grateful to all the families that participated in this study as well as to support charities Max Appeal, The 22Crew and Unique for their help and support. We thank all members of the IMAGINE-ID consortium for their contributions.

## Supplementary Methods

### Initial Variable Filtering

The initial dataset contained 1450 variables with information from 589 individuals (441 individuals with a ND-GC and 148 unaffected control individuals). To prepare the data for analysis, we began by removing those variables that contained administrative, free text and date and time information, as well as variables that were not quantitative questionnaire responses or coding of symptom intensity. Following these initial steps, variables where the most common response made up greater than 95% of responses to the question were removed as these items would likely not be useful in distinguishing young people with ND- GCs and phenotypic difficulties from other young people. In addition, those variables with a missing rate greater than 25% were also removed. Once the variables had been filtered, individuals with missing data rates across the remaining variables greater than 25% were also removed. These steps resulted in 489 individuals (376 ND-GC carriers [76.9%], of whom 41% had at least one sibling also included in the study) and 233 variables retained for further analysis.

### Principal *Components Analysis and Partial Least Squares Discriminant Analysis*

To develop an initial understanding of the dimensional structure of our data, we applied principal components analysis (PCA) followed by partial least squares discriminant analysis (PLSDA) to our training dataset, using the R *mixOmics* package (32). We used PCA as an initial unsupervised approach to identify the number of components that explained variance in our measured variables. Next, we applied a supervised approach (where the outcome was ND-GC status): sparse PLSDA. The number of components retained and number of variables per component were selected using 5-fold cross-validation, repeated 50 times, finding the combination that minimised prediction distance using one-sided t-tests testing for significant differences in the mean error rate when components are added to the model. The PLSDA model was then fit with the optimal number of components and variables.

### Model Evaluation

Elastic net regression models optimised penalty and mixture parameters; random forests used 1000 trees and optimised minimal node size and number of variables split at each node; SVM models optimised cost and margin parameters, neural network models optimised the number of hidden units, epochs and model penalty.

Model performance was evaluated for each outer fold by fitting the model with the best performing set of hyperparameters in the inner fold data to the (previously unseen) outer fold assessment dataset. This process was then repeated for all outer folds.

Following nested cross validation, we compared model performance based on the AUROC values for each outer fold, using a Bayesian linear mixed model fit with the R *rstanarm* package (52), where the outer fold identity was included as a varying intercept. From this model we calculated the performance of each model using the median of the posterior distribution, and the 95% credible interval using the highest density interval method. Models were then compared using the probability of direction method (53).

Modell variable importance was determined using permutation testing. This approach randomly permutes data from each variable in turn and evaluates the change in model performance (i.e., change in AUROC) following permutation. This was repeated 500 times to give a distribution of changes in performance after permutation. Variables with greater importance to the model will cause larger drops in AUROC than variables with lower importance

**Supplementary Table 1.** Counts of the genotypes of all study participants. *To preserve the confidentiality of individuals who had ND-GCs with a total count of < 5 participants with the same ND-GC in the study, we have grouped all such low frequency ND- GCs into a single group. This group contained 31 deletions and 25 duplications, with 15 other conditions being related to mixed deletions and duplications, single nucleotide variants, triplications, translocation, chromosomal trisomy, or imprinting. Chromosomal regions affected by ND-GCs in this group were: 1p21, 1p33, 1p36, 1q21, 1q42, 1q44, 2p12, 2p16, 2q11-q21, 2q13, 2q33, 2q34, 2q37, 3q28-29, 4p15, 4q28-31, 5p15, 5q23, 6p25, 6q27, 7p22, 7q11, 8q21, 8q24, 9p24, 9q34, 11q23, 12p13, 15pter-q13, 15q11, 15q11-q13, 15q13, 16p11, 16p12, 16p13, 16p21, 16q23, 17p11, 17p13, 17q12, 17q23, 17q25, 18p11, 20q13, 22q11, 22q12-q13, 22q13, Xp21, Xp22, Xp28.

| Genetic Condition | N |
| --- | --- |
| Controls | 104 |
| 16p11.2 proximal deletion | 45 |
| 15q11.2 deletion | 39 |
| 22q11.2 proximal deletion | 30 |
| 1q21.1 distal duplication | 28 |
| 15q13.3 deletion | 24 |
| 16p11.2 proximal duplication | 24 |
| 22q11.2 proximal duplication | 23 |
| 15q13.3 duplication | 20 |
| 1q21.1 distal deletion | 17 |
| NRXN1 deletion | 15 |
| 1q21.1 proximal TAR duplication | 13 |
| 16p11.2 distal deletion | 11 |
| Kleefstra Syndrome | 11 |
| 15q11.2 duplication | 5 |
| Other ND-GC* | 80 |

**Supplementary Table 2.**
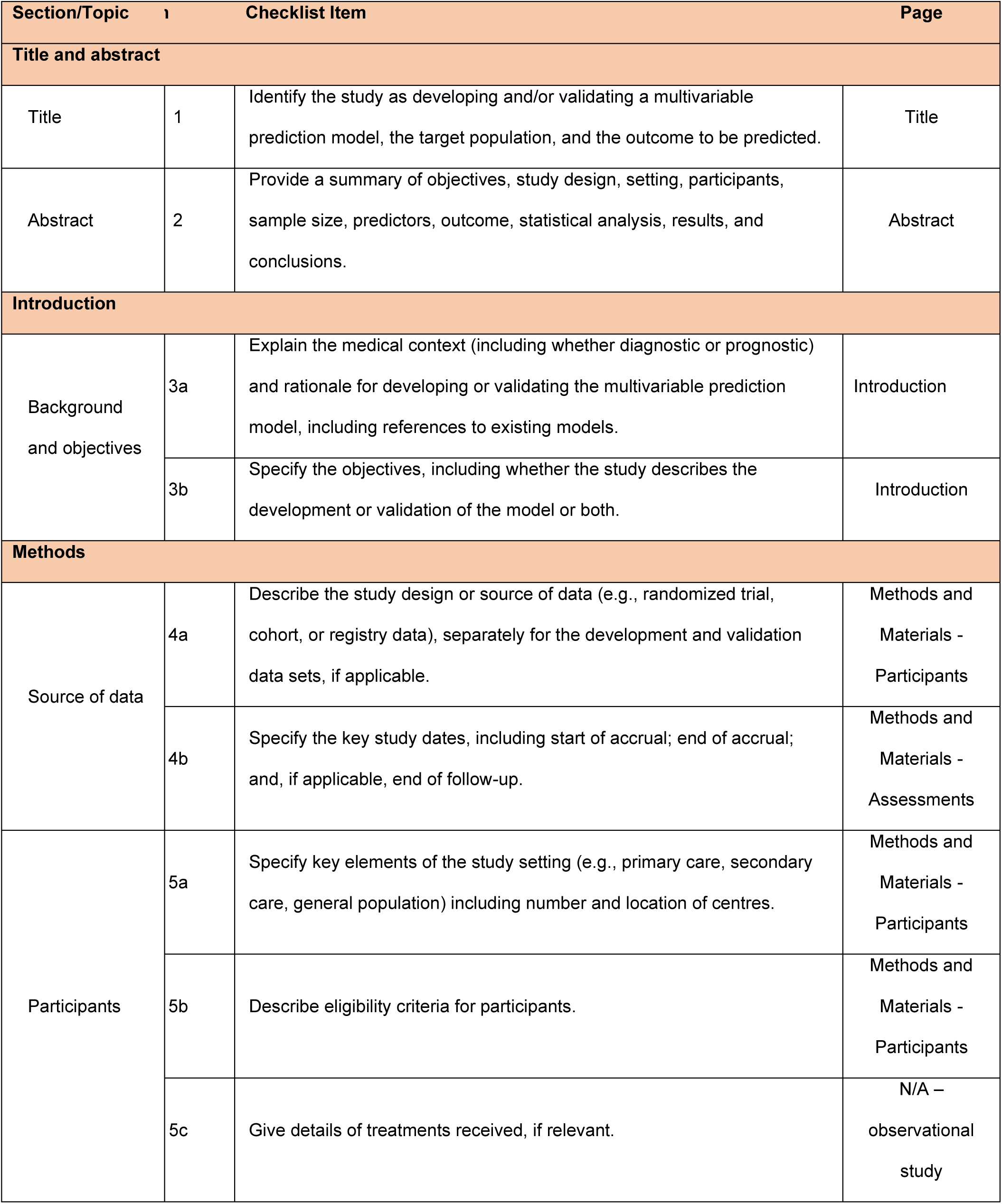
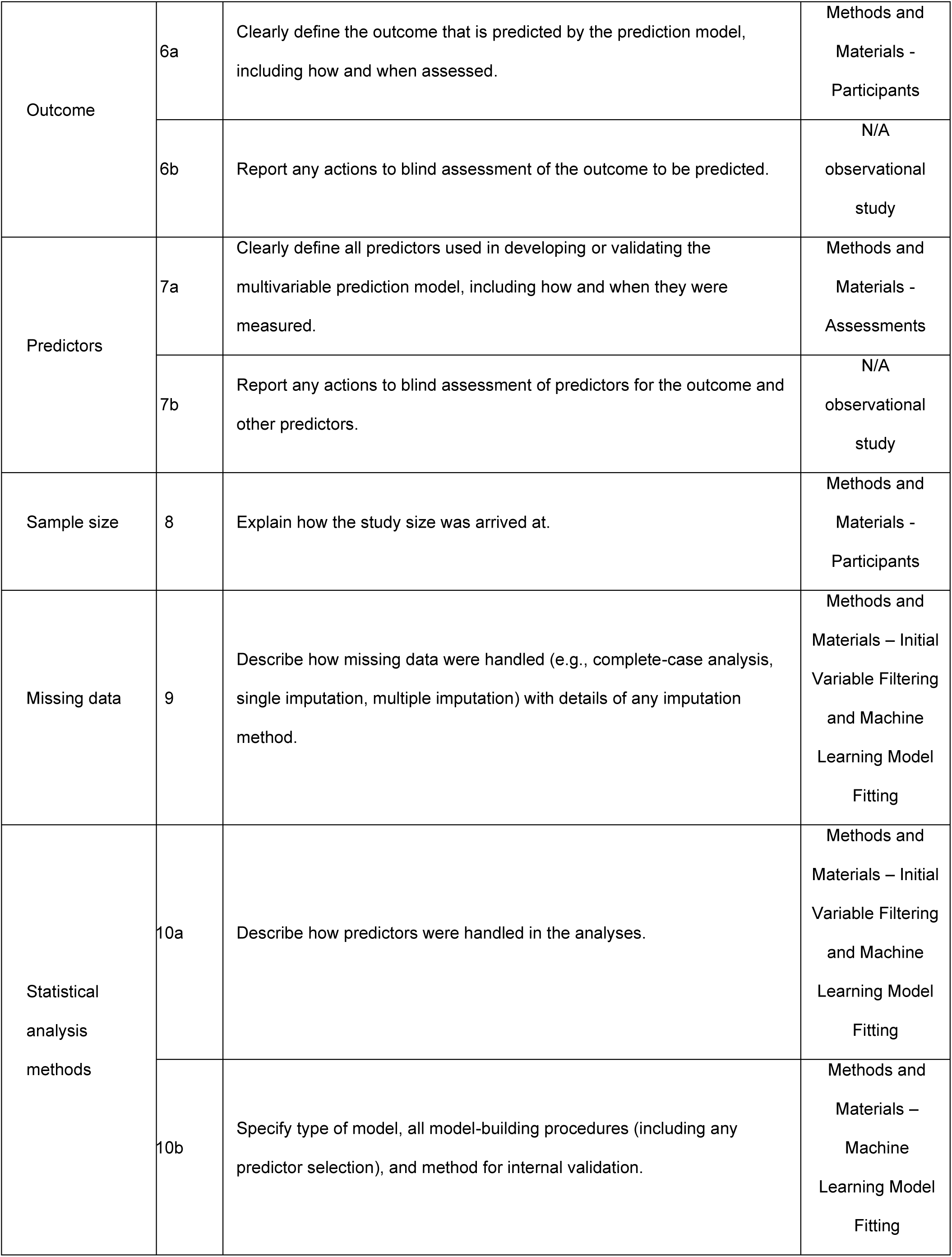
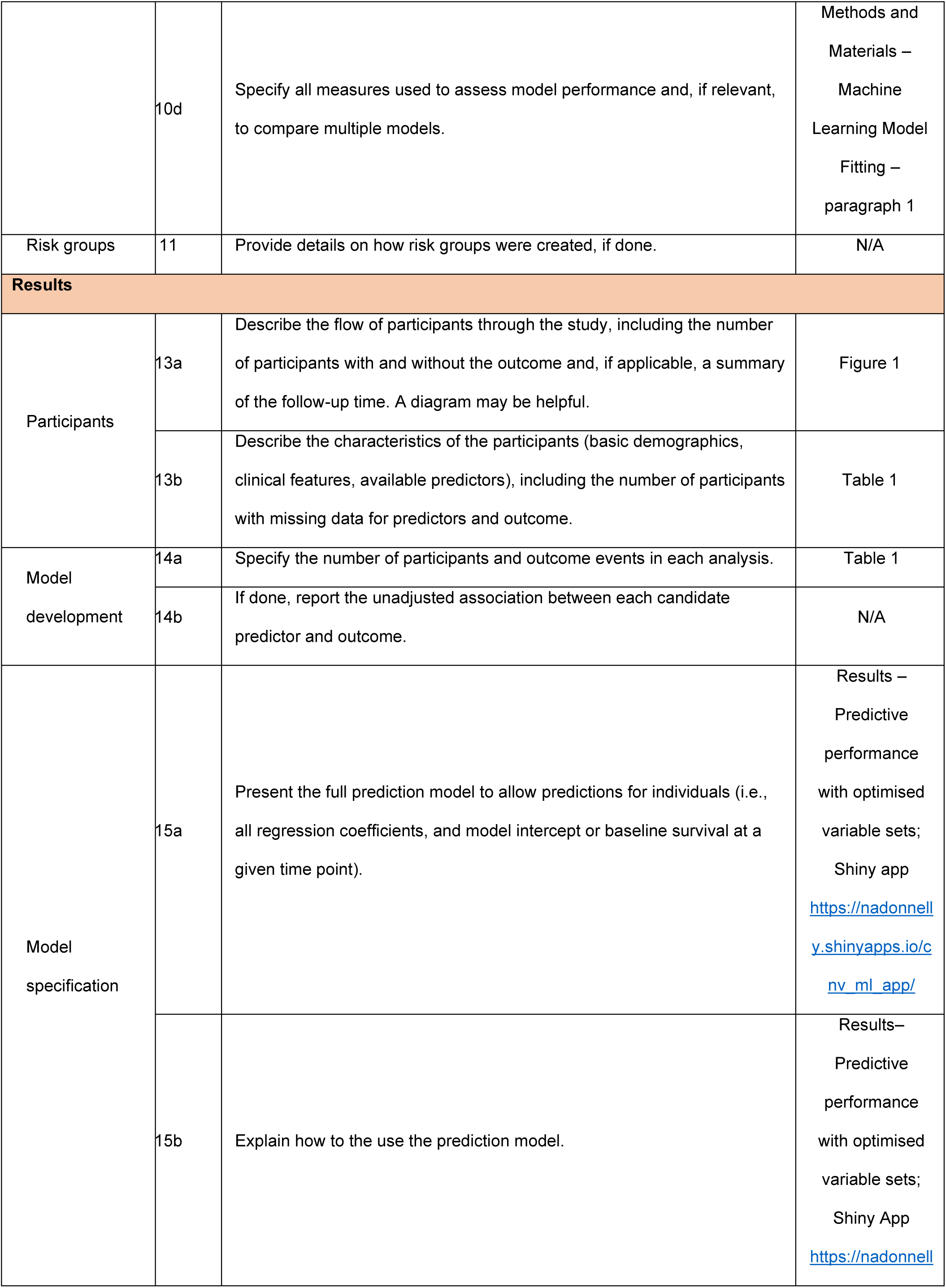
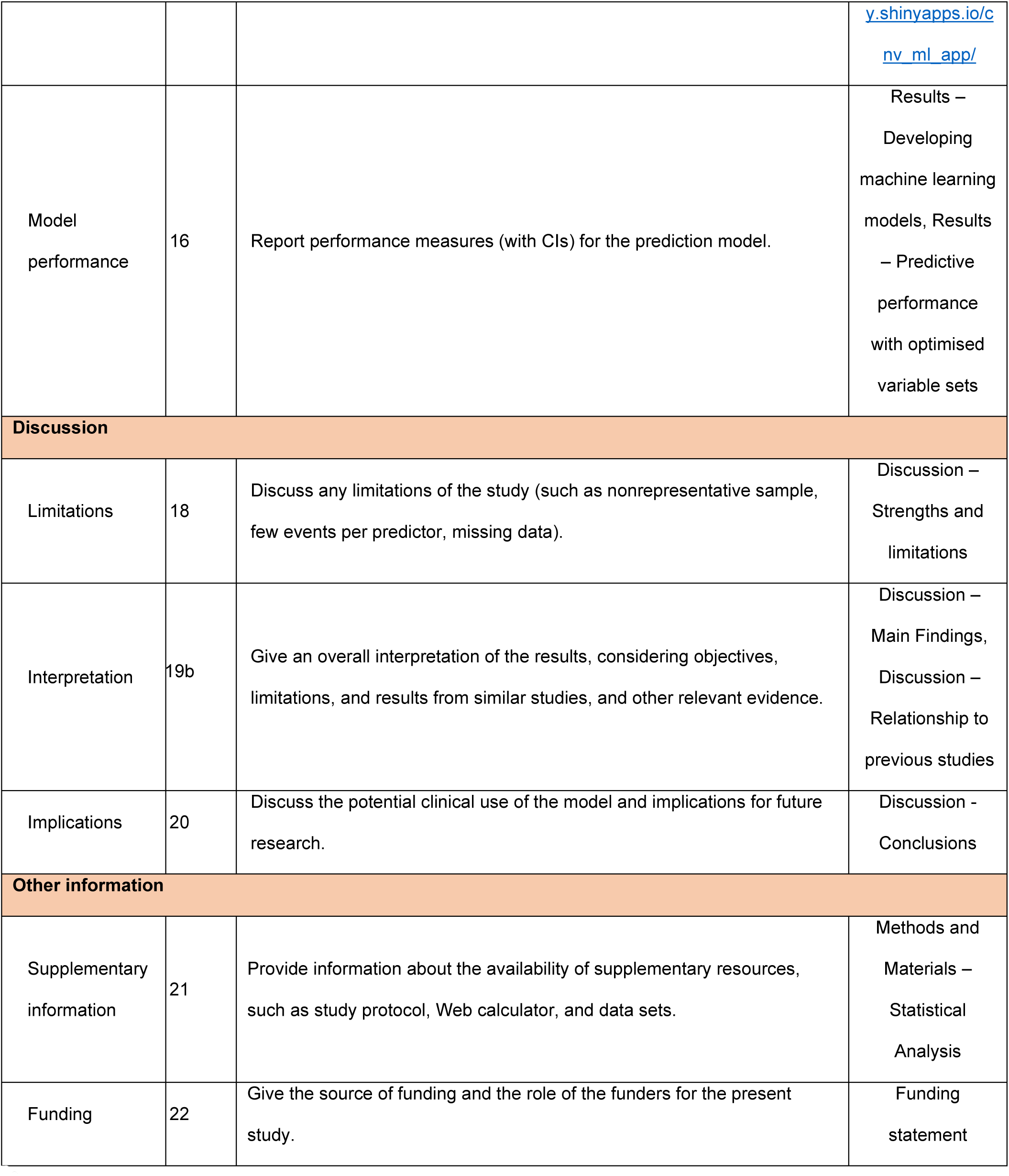
TRIPOD Reporting Guideline Table

**Supplementary Table 3.** Demographic information about the sample of children affected by a ND-GC and sibling controls.

| Variable |  | Group |  |  |
| --- | --- | --- | --- | --- |
|  | Overall,<br>N = 489<sup>1</sup> | Sibling<br>Control,<br>N = 113<sup>1</sup> | ND-GC,<br>N = 376<sup>1</sup> | p-value<sup>2</sup> |
| **Age** | 9.33<br>(7.27, 12.22) | 10.35<br>(8.13, 13.11) | 9.02<br>(7.12, 11.79) | <0.001 |
| **Gender** |  |  |  | 0.001 |
| Female | 180 (37%) | 56 (50%) | 124 (33%) |  |
| Male | 309 (63%) | 57 (50%) | 252 (67%) |  |
| **Highest Educational**<br>**Level** |  |  |  | 0.001 |
| No School Leaving Exams | 32 (6.5%) | 4 (3.5%) | 28 (7.4%) |  |
| Low | 102 (21%) | 22 (19%) | 80 (21%) |  |
| Middle | 174 (36%) | 38 (34%) | 136 (36%) |  |
| High | 128 (26%) | 25 (22%) | 103 (27%) |  |
| Unknown | 53 (11%) | 24 (21%) | 29 (7.7%) |  |
| **Income** |  |  |  | 0.009 |
| <=£19,999 | 123 (25%) | 21 (19%) | 102 (27%) |  |
| £20,000 - £39,999 | 164 (34%) | 35 (31%) | 129 (34%) |  |
| £40,000 - £59,999 | 73 (15%) | 14 (12%) | 59 (16%) |  |
| £60,000 + | 71 (15%) | 20 (18%) | 51 (14%) |  |
| Unknown | 58 (12%) | 23 (20%) | 35 (9.3%) |  |
| **Ethnicity** |  |  |  | <0.001 |
| European | 435 (89%) | 92 (81%) | 343 (91%) |  |
| Other | 31 (6.3%) | 5 (4.4%) | 26 (6.9%) |  |
| Unknown | 23 (4.7%) | 16 (14%) | 7 (1.9%) |  |
| <sup>1</sup> Median (IQR); n (%) |  |  |  |  |
| <sup>2</sup> Wilcoxon rank sum test; Pearson’s Chi-squared test |  |  |  |  |

**Supplementary Table 4.** Classification performance for each of the different machine learning techniques using training data and all variables. ANN: Artificial Neural Network; LR: Logistic Regression; SVM: Support Vector Machine

| Model | Performance | Difference | Probability of Direction |
| --- | --- | --- | --- |
| Linear SVM | 0.936 [0.917, 0.955] | - | 1 |
| Random Forest | 0.933 [0.915, 0.952] | -0.002 [-0.014, 0.008] | 0.654 |
| Penalised LR | 0.928 [0.909, 0.947] | -0.008 [-0.019, 0.003] | 0.172 |
| ANN | 0.9 [0.881, 0.919] | -0.036 [-0.047, -0.025] | 0 |

**Supplementary Table 5.** Classification performance for each of the different machine learning techniques using training data and different sets of variables. Column Performance is the median model performance over 20 outer folds of nested cross validation, estimated using a Bayesian generalised linear model, with 95% credible interval; Column Difference shows the model estimated difference in performance between the top performing model (Linear SVM with the top 30 model important variables estimated by the linear SVM fit to all variables) and a given model

| **Model** | **Variable Set** | **Performance** | **Difference** | **Probability**<br>**of Direction** |
| --- | --- | --- | --- | --- |
| **Linear SVM** | SVM | 0.944 [ 0.923,<br>0.964] | - | 1 |
| **Penalised LR** | SVM | 0.938 [ 0.918,<br>0.959] | -0.005 [ -0.021,<br>0.011] | 0.506 |
| **ANN** | SVM | 0.935 [ 0.915,<br>0.956] | -0.008 [ -0.025,<br>0.008] | 0.310 |
| **Random**<br>**Forest** | SVM | 0.942 [ 0.921,<br>0.962] | -0.002 [ -0.018,<br>0.014] | 0.814 |
| **Linear SVM** | Penalised LR | 0.942 [ 0.922,<br>0.963] | -0.002 [ -0.018,<br>0.014] | 0.812 |
| **Penalised LR** | Penalised LR | 0.935 [ 0.914,<br>0.955] | -0.009 [ -0.025,<br>0.007] | 0.300 |
| **ANN** | *Penalised LR* | *0.905 [ 0.885,*<br>*0.926]* | *-0.038 [ -0.054, -*<br>*0.022]* | 0 |
| **Random**<br>**Forest** | Penalised LR | 0.936 [ 0.917,<br>0.958] | -0.007 [ -0.024,<br>0.009] | 0.384 |
| **Linear SVM** | ANN | *0.925 [ 0.904,*<br>*0.945]* | *-0.019 [ -0.035, -*<br>*0.002]* | 0.022 |
| **Penalised LR** | ANN | *0.926 [ 0.905,*<br>*0.946]* | *-0.018 [ -0.034, -*<br>*0.002]* | 0.030 |
| **ANN** | ANN | *0.922 [ 0.901,*<br>*0.942]* | *-0.022 [ -0.038, -*<br>*0.006]* | 0.008 |
| **Random Forest** | ANN | 0.937 [ 0.916, 0.957] | -0.007 [ -0.023, 0.009] | 0.418 |
| **Linear SVM** | Random Forest | 0.938 [ 0.918, 0.959] | -0.006 [ -0.022, 0.011] | 0.492 |
| **Penalised LR** | Random Forest | 0.933 [ 0.912, 0.953] | -0.011 [ -0.027, 0.005] | 0.184 |
| **ANN** | *Random Forest* | *0.917 [ 0.896, 0.938]* | *-0.027 [ -0.043, -0.011]* | *0* |
| **Random Forest** | Random Forest | 0.942 [ 0.921, 0.962] | -0.002 [ -0.018, 0.015] | 0.842 |
| **Linear SVM** | All Variables | 0.933 [ 0.914, 0.955] | -0.01 [ -0.027, 0.006] | 0.214 |
| **Penalised LR** | All Variables | 0.926 [ 0.905, 0.946] | -0.018 [ -0.034, -0.002] | 0.028 |
| **ANN** | *All Variables* | *0.898 [ 0.877, 0.919]* | *-0.046 [ -0.062, -0.03]* | *0* |
| **Random Forest** | All Variables | 0.931 [ 0.91, 0.951] | -0.013 [ -0.029, 0.003] | 0.120 |
| **Linear SVM** | Variables selected by > 1 model | 0.944 [ 0.923, 0.964] | 0 [ -0.016, 0.016] | 0.986 |
| **Penalised LR** | Variables selected by > 1 model | 0.936 [ 0.915, 0.956] | -0.008 [ -0.023, 0.009] | 0.350 |
| **ANN** | Variables<br>selected by ><br>1 model | 0.923 [ 0.902,<br>0.943] | -0.021 [ -0.037, -<br>0.005] | 0.012 |
| **Random**<br>**Forest** | Variables<br>selected by ><br>1 model | 0.938 [ 0.918,<br>0.959] | -0.006 [ -0.022,<br>0.01] | 0.498 |
| **Linear SVM** | Variables<br>selected by ><br>2 models | 0.944 [ 0.923,<br>0.964] | 0 [ -0.017, 0.016] | 0.982 |
| **Penalised LR** | Variables<br>selected by ><br>2 models | 0.94 [ 0.92,<br>0.962] | -0.003 [ -0.019,<br>0.013] | 0.704 |
| **ANN** | Variables<br>selected by ><br>2 models | 0.934 [ 0.914,<br>0.955] | -0.01 [ -0.026,<br>0.006] | 0.234 |
| **Random**<br>**Forest** | Variables<br>selected by ><br>2 models | 0.939 [ 0.918,<br>0.959] | -0.005 [ -0.021,<br>0.011] | 0.568 |

**Supplementary Table 6:** Performance statistics split by age and gender

| Covariate | Value | AUROC | Mean Log Loss |
| --- | --- | --- | --- |
| Age (quintile) | [5.89,6.72] | 1 | 0.105 |
|  | (6.72,8.57] | 0.964 | 0.288 |
|  | (8.57,9.41] | 1 | 0.28 |
|  | (9.41,12.2] | 1 | 0.108 |
|  | (12.2,21.6] | 0.987 | 0.201 |
| Gender | Female | 0.949 | 0.312 |
|  | Male | 0.99 | 0.132 |

**Supplementary Table 7.** Final variables and associated dimensions identified using bootstrap exploratory graph analysis.

| Variable Name | Variable Definition | Dimension |
| --- | --- | --- |
| SAP | Separation Anxiety: do they get aches and pains, feel sick, get headaches etc. on school days, or at other times when separated from a parent/carer (Binary Variable) | 1: Anxiety |
| SAI | Separation Anxiety: Excessive worries or fear concerning separation from the persons to whom the child is attached (Binary Variable) | 1: Anxiety |
| FPB | Fear of activities in public: does fear of activities in public lead to a restricted lifestyle (Binary Variable) | 1: Anxiety |
| SAS | Separation Anxiety: Avoidance, or attempted avoidance, of sleeping away from family, as a result of worrying or anxiety about separation from home or family (Binary Variable) | 1: Anxiety |
| SAF | Separation Anxiety: The child sleeps with a family member because of persistent refusal to sleep through the night without being near a major attachment figure (Binary Variable) | 1: Anxiety |
| AGO | Agoraphobia: Does agoraphobia lead to a restricted lifestyle (Binary Variable) | 1: Anxiety |
| EST | Health and Development: Is your child educationally statemented? (Binary Variable) | 2: Development/<br>Co-ordination |
| SLT | Health and Development: Has your child had speech therapy? (Binary Variable) | 2: Development/<br>Co-ordination |
| W18 | Health and Development: Did your child walk by 18 months of age? (Binary Variable) | 2: Development/<br>Co-ordination |
| CBA | Coordination and motor development: Your child catches a small ball (e.g. tennis ball size) thrown from a distance of 6-8 feet (1.8-2.4 metres) (Ordinal variable: 1: Not at all like your child; 2: A bit like your child; 3: Moderately like your child; 4: Quite a bit like your child; 5: Extremely like your child) | 2: Development/<br>Co-ordination |
| CRF | Coordination and motor development: Your child runs as fast and in a similar way to other children of the same age and gender (Ordinal variable: 1: Not at all like your child; 2: A bit like your child; 3: Moderately like your child; 4: Quite a bit like your child; 5: Extremely like your child) | 2: Development/<br>Co-ordination |
| COB | Coordination and motor development: If your child has a plan to do a motor activity, they can organise their body to follow the plan and effectively complete the task (e.g., building a cardboard or cushion 'fort', moving on playground equipment, building a house or a structure with blocks, or using craft materials) (Ordinal variable: 1: Not at all like your child; 2: A bit like your child; 3: Moderately like your child; 4: Quite a bit like your child; 5: Extremely like your child) | 2: Development/<br>Co-ordination |
| CCP | Coordination and motor development: Your child cuts pictures and shapes accurately (Ordinal variable: 1: Not at all like your child; 2: A bit like your child; 3: Moderately like your child; 4: Quite a bit like your child; 5: Extremely like your child) | 2: Development/<br>Co-ordination |
| REA | Health and Development: Is your child behind in reading? (Binary Variable) | 2: Development/<br>Co-ordination |
| MSK | Health and Development: Has your child had skeletal or muscular problems? (Binary Variable) | 2: Development/<br>Co-ordination |
| InT | Insomnia: Does the child experience overall insomnia greater than 1 hour per night? (Binary Variable) | 3: Insomnia |
| InI | Insomnia: Is initial insomnia present (Does it take more than an hour to get to sleep at night?) (Binary Variable) | 3: Insomnia |
| DeI | Depression: Was there a week when the participant felt miserable most days? (Binary Variable) | 4: Depression |
| D2M | Depression: Has there been a period of two months in the last year when the participant did not feel depressed in mood? (Binary Variable) | 4: Depression |
| DeQ | Depression: Depressed mood has a subjectively different quality from sadness, contrasted with an experience that caused sadness, such as loss of a pet or watching a sad film (Binary variable) | 4: Depression |

